# Beat-to-Beat Action Potential Duration Variability, Not Duration, Reflects β-Adrenergic Stimulation-Induced Repolarization Instability

**DOI:** 10.64898/2026.10.02.26364548

**Authors:** Daisuke Sato, Roshni Shetty, Raffaele Coppini, Eleonora Grandi

## Abstract

Instability of the cardiac electrical system is a major cause of life-threatening arrhythmias. Current safety guidelines have traditionally emphasized QT interval prolongation on the electrocardiogram (ECG) as a primary biomarker of proarrhythmic risk. However, this approach does not capture all forms of electrical instability. In this study, we demonstrate that beat-to-beat variability in action potential duration (APD), a cellular surrogate of the QT interval, better stratifies proarrhythmic risk than mean APD prolongation alone. Using a validated computational model of a human ventricular myocyte, we simulated the effects of the β-adrenergic agonist isoproterenol (ISO). Increasing the ISO concentration shortened mean APD but paradoxically increased beat-to-beat APD variability, measured as the standard deviation of APDs. In our simulations, the emergence of early afterdepolarizations (EADs), abnormal oscillations of the membrane potential during the plateau phase of the action potential, at higher ISO concentrations was consistently preceded by a marked rise in APD variability, even while mean APD was reduced. Mechanistic analysis revealed that ISO-induced enhancement of L-type Ca^2+^ current (I_CaL_) and slow delayed rectifier K^+^ current (I_Ks_) steepens the APD restitution slope, which reflects action potential instability, even when the net effect is a reduction in APD. These results support APD variability as a more sensitive and mechanistically plausible marker of proarrhythmic risk than APD prolongation alone. Our findings suggest that QT interval variability may improve risk stratification and provide a more robust mechanistic foundation for drug safety evaluation.

**Significance Statement:** QT interval prolongation remains an important component of cardiac safety assessment, but changes in mean QT duration may not fully capture the dynamic stability of ventricular repolarization. Here, we show that beat-to-beat APD variability increases during β-adrenergic stimulation even when mean APD shortens, and increases further as the system approaches conditions associated with EAD formation. These findings identify APD variability as a complementary measure of repolarization stability and provide a cellular mechanistic basis for further evaluating QT interval variability as an additional marker of proarrhythmic susceptibility.

## Introduction

Sudden cardiac death (SCD), a major global health challenge, is often caused by lethal ventricular tachyarrhythmias such as Torsades de Pointes (TdP) (1,2). A critical and preventable trigger for TdP is drug-induced proarrhythmia (3,4), which extends well beyond cardiovascular drugs to many commonly prescribed medications such as antibiotics, antipsychotics, and antihistamines (5-7). The risk that new therapeutic agents may induce TdP is a critical safety concern during drug development, and cardiotoxicity remains a leading cause of both drug withdrawal and termination of candidate compounds. These realities create an immense clinical and economic imperative to establish accurate and mechanistically robust methodologies for assessing proarrhythmic risk.

Current regulatory practice is guided by the International Council for Harmonisation (ICH) S7B and E14 guidelines, which rely on hERG channel block potency and QT interval prolongation on the electrocardiogram (ECG) as primary biomarkers of TdP risk (8-11). More recent regulatory efforts, including the updated ICH E14/S7B framework and the Comprehensive in vitro Proarrhythmia Assay (CiPA), have expanded this paradigm toward integrated, mechanistically informed assessment incorporating multichannel pharmacology and computational modeling (11-14). Nevertheless, QT/QTc prolongation remains an important clinical indicator of delayed ventricular repolarization and continues to play a central role in cardiac safety evaluation. This is based on the observation that most torsadogenic drugs are potent hERG blockers. By inhibiting the rapid component of the delayed rectifier potassium current (I_Kr_), these drugs prolong the cellular action potential duration (APD) and consequently the QT interval. Although this QT prolongation-based framework has successfully prevented many high□risk drugs from reaching the market, clinical and experimental studies have revealed major limitations of QT prolongation as a standalone biomarker. QT prolongation lacks specificity, failing to distinguish between drugs causing benign QT prolongation and those posing genuine TdP risk. For example, amiodarone markedly prolongs the QT interval and is associated with a known risk of TdP, but with a low incidence (15). Similarly, ranolazine prolongs the QT interval yet it is only associated with TdP when administered to patients with hypokalemia, hypomagnesemia, or bradycardia, or when co-administered with drugs that either prolong the QT interval or inhibit the metabolic breakdown of ranolazine (16). The current QT-based framework can thus both misclassify safe drugs as hazardous, leading to premature termination of valuable therapeutics (false positives), and potentially overlook compounds destabilizing repolarization without prolongation (false negatives). The disconnect between QT prolongation and true arrhythmic liability highlights the need for biomarkers that more directly capture the electrophysiological processes that initiate and sustain TdP.

An emerging concept is that the dynamic stability of repolarization, rather than its duration alone, may more accurately reflect proarrhythmic vulnerability. Our recent studies have shown that beat-to-beat variability of APD at the cellular level, and QT interval variability (QTV) on the ECG, capture this instability (17-19). In this study, we test the hypothesis that APD variability is a more robust and reliable predictor of cellular arrhythmogenesis than APD prolongation alone. Although many torsadogenic agents prolong APD, proarrhythmic instability can emerge in the absence of appreciable APD prolongation. β-adrenergic stimulation provides a particularly relevant example, as it shortens APD while increasing repolarization instability and promoting afterdepolarizations (20-22). These dual effects make isoproterenol an ideal pharmacological test case for separating APD prolongation from its beat-to-beat variability and dynamic instability. This work establishes mechanistic links between APD variability, QTV, and proarrhythmic events, providing a biophysically grounded framework for arrhythmic risk assessment.

## Methods

### Human Ventricular Myocyte Action Potential Model

To investigate the cellular electrophysiological response to β-adrenergic stimulation, we used the human ventricular myocyte model developed by Shetty et al. (2025) (23) as our starting point. The model is based on the physiologically detailed O’Hara-Rudy ventricular cell model of the human ventricular endocardial myocyte (24) and incorporate previously established modifications to improve its representation of human ventricular electrophysiology. Specifically, the original *I*_Na_ formulation was replaced with the modified Ten Tusscher formulation, as implemented in the Grandi et al. (2010) ventricular myocyte model (25) to improve conduction behavior in tissue simulations. Sex-specific differences were incorporated and parameterized to reproduce male and female electrophysiological phenotypes as described in Shetty et al. (2025) (23). Additionally, conductances governing *I*_Kr_, *I*_Ks_, *I*_K1_, *I*_CaL_, and *I*_NaL_ were rescaled following the modifications reported by Dutta et al. 2017 (26) to reproduce the optimized human ventricular electrophysiological phenotype used for proarrhythmia studies.

### Signaling Pathway and Simulation of β-Adrenergic Stimulation with Isoproterenol (ISO)

β-Adrenergic regulation was incorporated following the framework of Gong *et al*., 2021, which couples compartmentalized β-adrenergic signaling to human ventricular electrophysiology. The signaling network was adopted from the Heijman et al. 2011 canine model (27). The model describes ISO binding to β□- and β□-adrenergic receptors, activation of adenylyl cyclase, and subsequent cAMP production leading to protein kinase A (PKA) activation. The activated PKA catalytic subunit modulates electrophysiology and Ca^2^□ handling through phosphorylation of targets distributed across three cellular compartments: (1) the caveolar domain (CAV), including L-type Ca^2^□ channels, fast Na□ channels, Na□/K□ pumps, and ryanodine receptors; (2) the extracaveolar domain (ECAV), containing slow delayed rectifier and background K□ channels; and (3) the cytosolic domain (CYT), including phospholamban (PLB) and troponin I (TnI). The phosphorylated fraction of each target is treated as a dynamic state variable determined by local β-AR signaling at each ISO concentration (0–1.0 µM), where 1.0 µM produces maximal β-adrenergic stimulation.

### Stochastic Simulation and Quantification of APD Variability

Stochastic fluctuations were incorporated as previously described in Sato et al. (19) to reproduce physiological beat-to-beat variability in ventricular repolarization. Stochastic channel gating was implemented using a Langevin formulation applied to the gating variable of *I*_KS_ . The effective population of *I*_KS_ channels was set to 10,000. For each condition, the model was pre-paced for 1,000 beats to reach steady state, after which it was paced at the indicated pacing cycle length (PCL), and the APD was recorded over 100,000 consecutive beats. APD was defined as the time interval between the depolarization and repolarization threshold crossings at a membrane potential of -80 mV. Beat-to-beat APD variability was quantified as the standard deviation of APD across these beats.

### Numerical Simulation Protocols and Code Availability

All codes were written and implemented in C++. The ordinary differential equations were solved using the fourth-order Runge-Kutta method with a time step of 0.005 ms. The source code is freely accessible on our websites (GitHub / Zenodo).

## Results

### β-adrenergic stimulation markedly increases beat-to-beat variability of APD despite APD shortening

We first quantified the effect of β-adrenergic stimulation on the APD. The female model was selected as the primary testbed because female ventricular electrophysiology is associated with greater susceptibility to repolarization instability and acquired proarrhythmia, providing a sensitive substrate to test whether APD variability can detect instability that is not captured by mean APD alone. As shown in **Fig. 1A**, increasing concentrations of ISO induced a progressive shortening of the APD. At baseline (0 nM ISO) and a pacing cycle length (PCL) of 2000 ms, the APD was 366.4 ms. ISO shortened APD in a concentration-dependent manner, reaching a minimum of 308.7 ms at 40 nM, corresponding to a maximal decrease of ∼15.7%. Further increasing [ISO] eventually produced APD prolongation with the emergence of EADs at ∼60 nM ISO, which occurred only during slow pacing (e.g., PCL = 2000 ms). These EADs are driven by the reactivation of the LTCCs. The observed APD shortening is consistent with experimental findings and is primarily attributed to the dominant effect of ISO to enhance I_Ks_ (21,28) over I_CaL_ increase.

**Figure 1.**
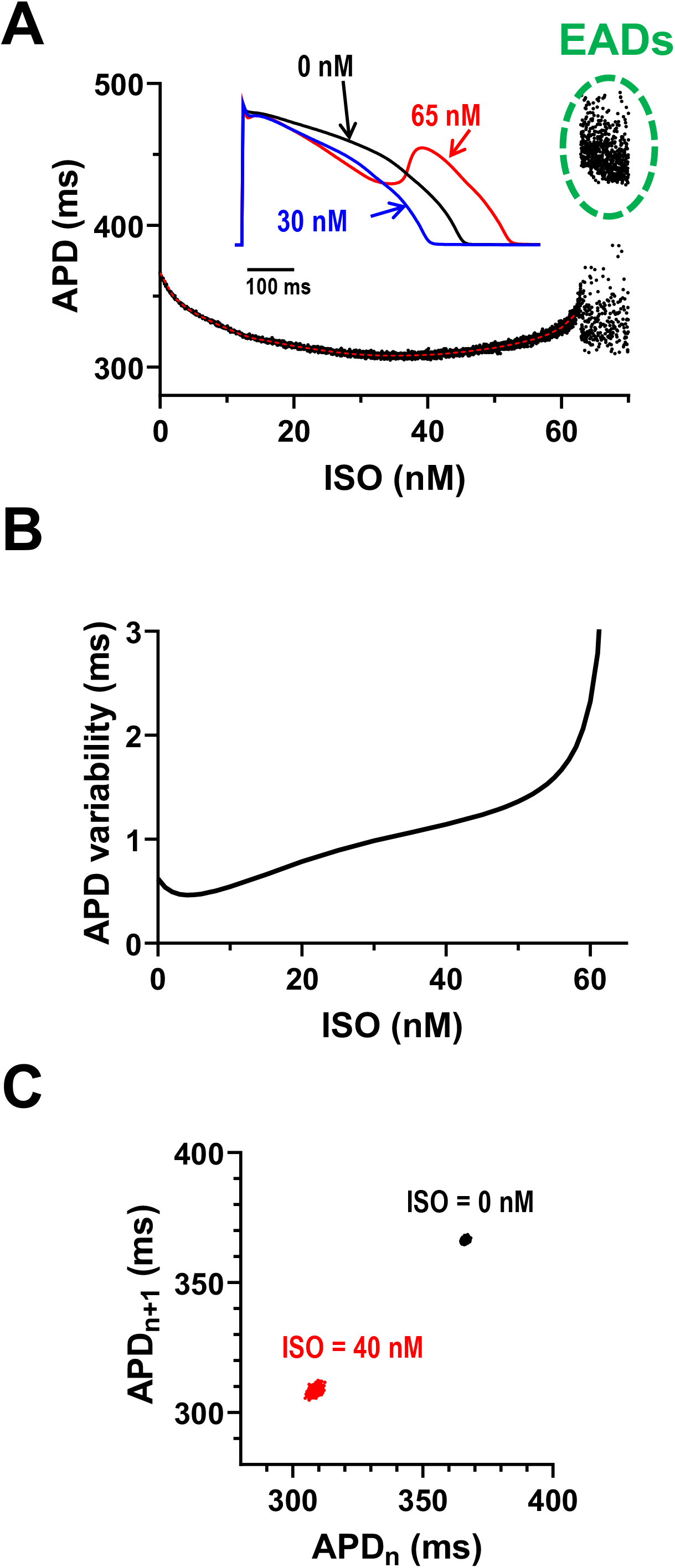
Effects of ISO on APD and APD Variability. **(A)** APD as a function of ISO concentration. The red dashed line indicates the mean APD. Inset: representative action potentials at baseline (black), ISO = 30 nM (blue) and 65 nM (red). **(B)** Beat-to-beat APD variability (measured as standard deviation) as a function of ISO concentration. **(C)** Poincaré plots (APD_n+1_ vs. APD_n_) for 1,000 consecutive beats at baseline (black dots, tight cluster) and in the presence of 40 nM ISO (red dots, diffuse cloud). All simulations were performed at PCL = 2000 ms.

In contrast to shortening the APD, β-adrenergic stimulation induced a substantial increase in beat-to-beat APD variability. **Fig. 1B** shows the concentration-dependent effect of ISO on APD variability. At baseline, the action potentials were highly stable, with an APD variability of 0.62 ms. At ISO = 4 nM, APD variability reached a minimum of 0.46 ms. As the ISO concentration was increased, APD variability increased, reaching 2.33 ms at 60 nM. This observed increase in APD variability is consistent with experimental reports of increased QTV under ISO stimulation (29). This increase in APD variability and the shortening of APD are visualized in the Poincaré plot shown in **Fig. 1C**. At baseline, the plot of APD_n+1_ vs. APD_n_ for 1,000 consecutive beats shows a tight cluster of points, an indication of highly periodic behavior. In the presence of 40 nM ISO, the same plot reveals a more widely scattered, diffuse cloud of points, reflecting larger beat-to-beat APD fluctuations despite shorter APD values.

### APD variability parallels restitution steepening, not mean APD

We next assessed dynamical instability by calculating the slope of the APD restitution curve. It is known that ISO steepens the APD restitution slope (30,31). **Fig. 2A** shows the S1-S2 APD restitution curves when S1 was 2000 ms, with steeper slopes indicating greater instability. The restitution slope increased as a function of ISO concentration (**Fig. 2B**) following a pattern that closely parallels APD variability (**Fig. 1B**) rather than the mean APD (**Fig. 1A**). This relationship is further illustrated in **Fig. 2C**, where APD variability increases approximately linearly with restitution slope. This suggests that APD variability tracks dynamical instability more closely than mean APD.

**Figure 2.**
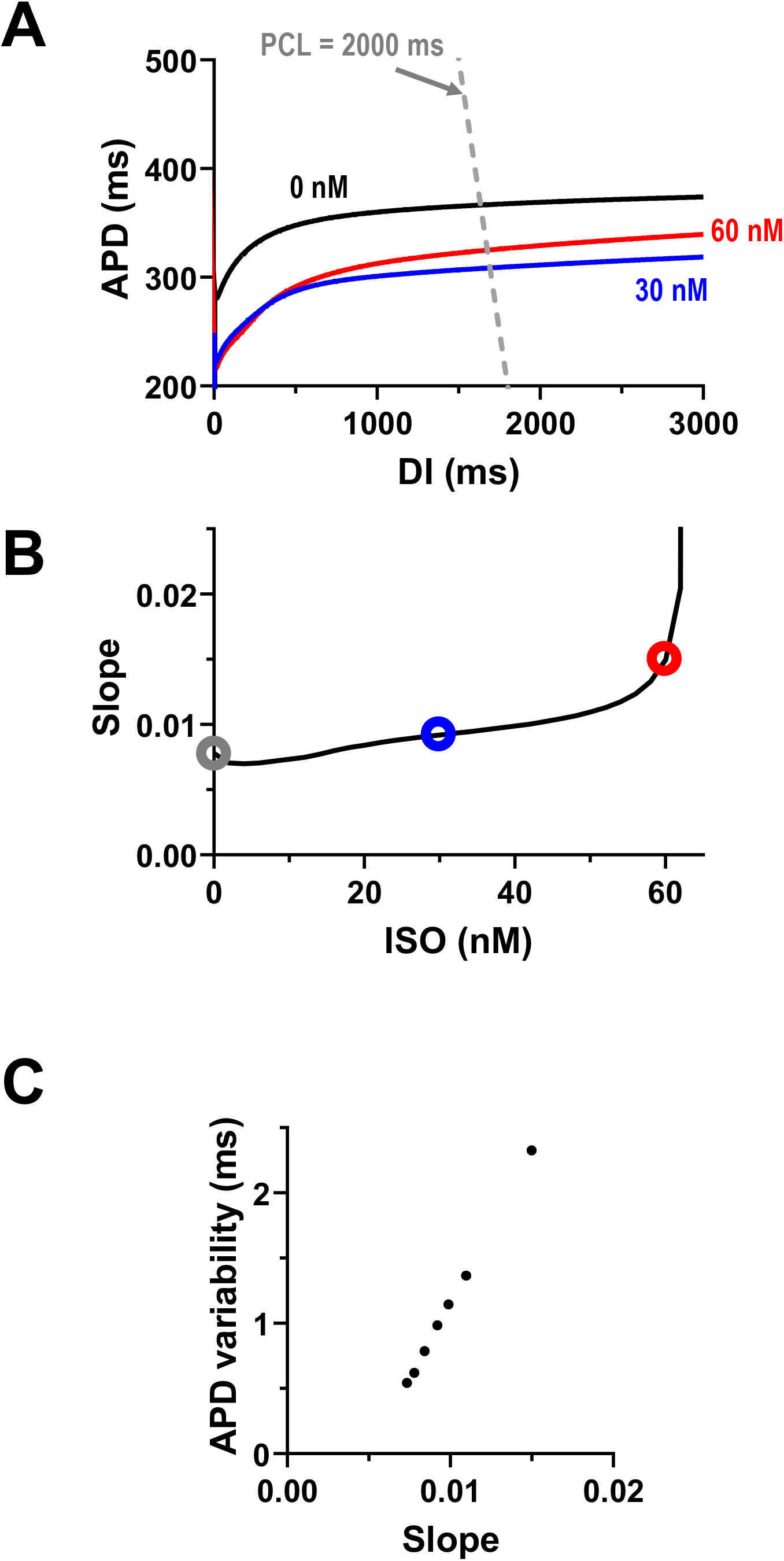
APD Variability is Linearly Correlated with Dynamical Instability. **(A)** S1-S2 APD restitution curves for different ISO concentrations. S1 = 2000 ms. The gray dashed line indicates PCL = 2000 ms (PCL = APD + DI). The restitution slope is evaluated at the intersection points of this line and the restitution curves. **(B)** APD restitution slope (at PCL = 2000 ms) as a function of ISO concentration. The pattern parallels APD variability (Fig. 1B). **(C)** APD variability vs. the restitution slope.

To further examine whether APD variability reflects changes in repolarization dynamics independently of mean APD, we selected two ISO concentrations (16 and 55 nM) that yielded the same APD (**Fig. 3A**). While ISO enhances both I_CaL_ and I_Ks_ in a dose-dependent manner, these opposing effects counterbalance each other and maintain the APD unchanged (**Fig. 3B**). However, despite the unchanged mean APD, beat-to-beat variability is noticeably larger at 55 nM compared to 16 nM, as visualized in the Poincaré plots (**Fig. 3C**). We then tested the changes in APD variability when APD is held constant. As ISO was varied, I_Ks_ (G_Ks_) was adjusted so that APD remained equal to the baseline value of 366.4 ms. **Fig. 4A** shows the required G_Ks_ adjustments (relative to the ISO-modified G_Ks_) across ISO concentrations. As a result, APD remained constant across ISO concentrations from 0 to 60 nM (**Fig. 4B**). However, APD variability continued to rise steadily with [ISO] (>6 nM), and concentrations above approximately 60 nM precipitated AP instability and EADs (**Fig. S1**). We also constructed S1-S2 restitution curves under these conditions (**Fig. 5A** and **Fig. 5B**). Although the restitution curves intersect at PCL = 2000 ms having the same APD at this pacing rate, their local slopes differ, indicating different degrees of dynamical instability. When the restitution slope is plotted as a function of ISO concentration (**Fig. 5C**), its profile closely parallels the APD variability pattern shown in **Fig. 4C**. Together, these results demonstrate a clear link between APD variability and dynamical instability, and a dissociation between mean APD and the underlying proarrhythmic instability. Qualitatively similar results were obtained when APD was held constant by adjusting I_CaL_ instead of I_Ks_, confirming that the observed relationship between APD variability and dynamical instability is robust and independent of the specific ionic mechanism used to preserve APD (**Fig. S2**).

**Figure 3.**
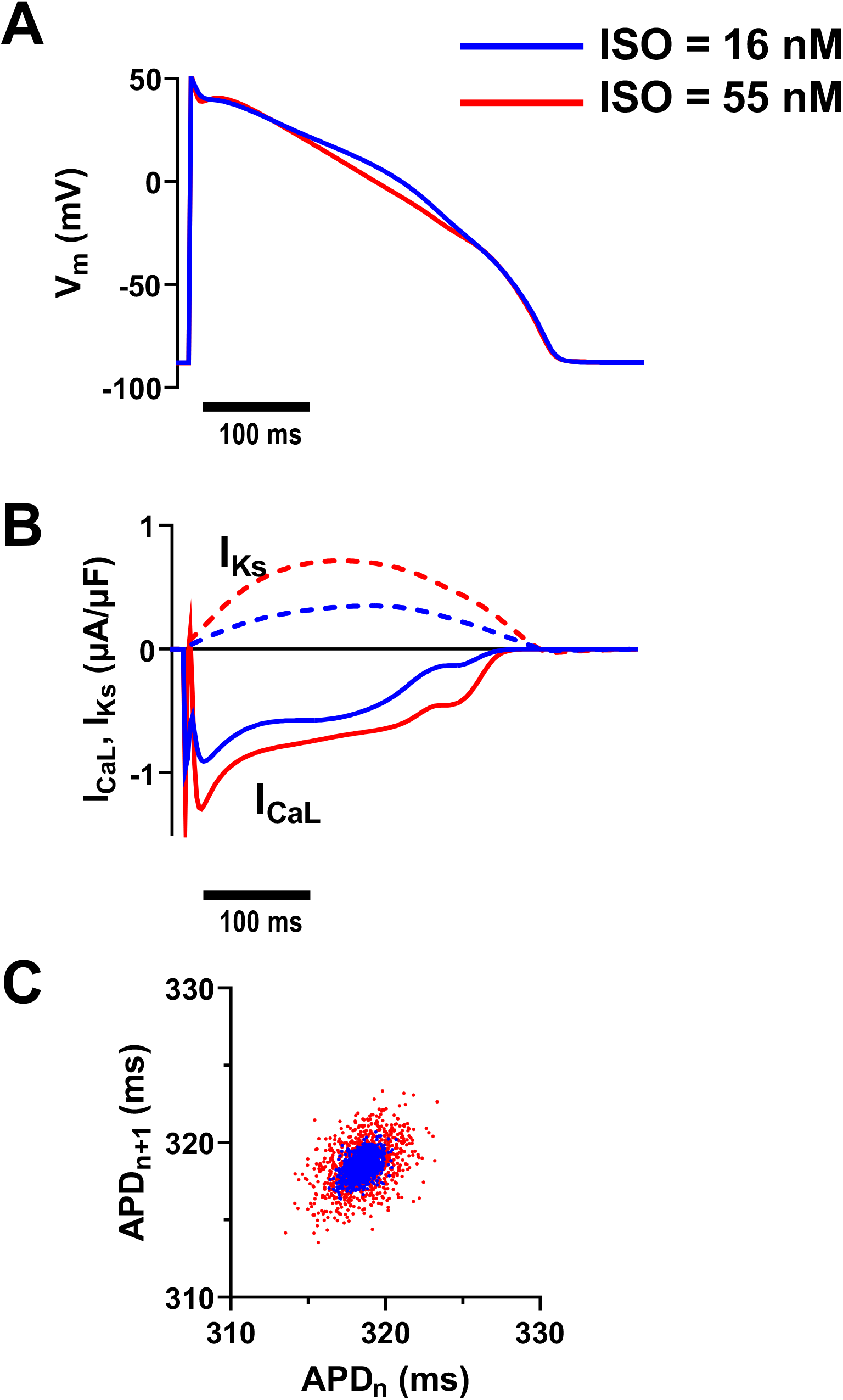
Time Course of Action Potential and Underlying Currents. **(A)** Membrane voltage vs. time at ISO = 16 nM (blue) and ISO = 55 nM (red). **(B)** I_CaL_ (solid lines) and I_Ks_ (dashed lines) vs. time. **(C)** Poincaré plots (APD_n+1_ vs. APD_n_) for 1,000 consecutive beats at ISO = 16 nM (blue dots, tight cluster) and ISO = 55 nM (red dots, diffuse cloud).

**Figure 4.**
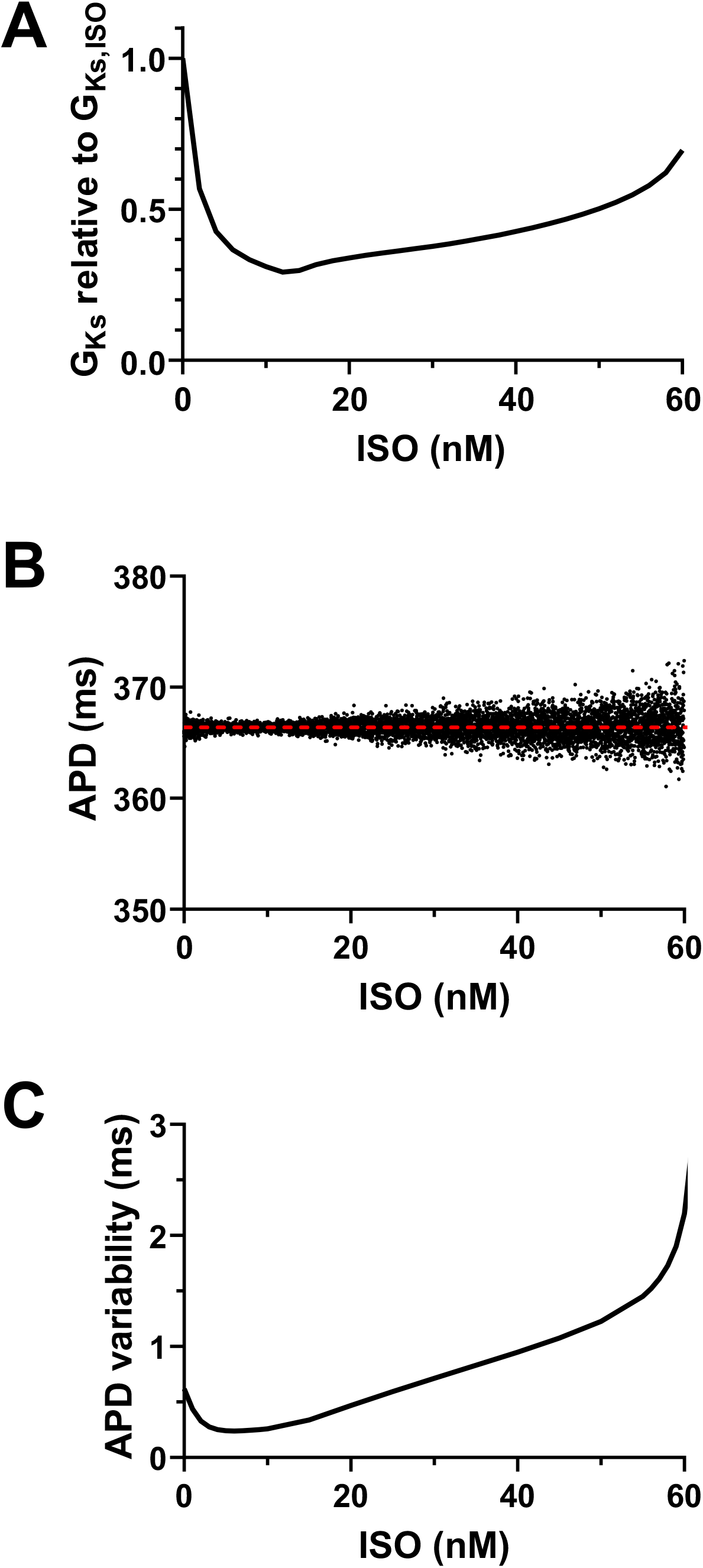
Constant Mean APD Conditions. **(A)** Required adjustment factor for G_Ks_ (relative to the ISO-modified G_Ks_) to maintain a constant APD of 366.4 ms across all ISO concentrations. G_Ks,adjusted_ = (adjustment factor) × G_Ks,ISO_ **(B)** APD as a function of ISO concentration under G_Ks_ adjustment. The dashed line indicates the mean APD. **(C)** APD variability as a function of ISO concentration under constant mean APD conditions.

**Figure 5.**
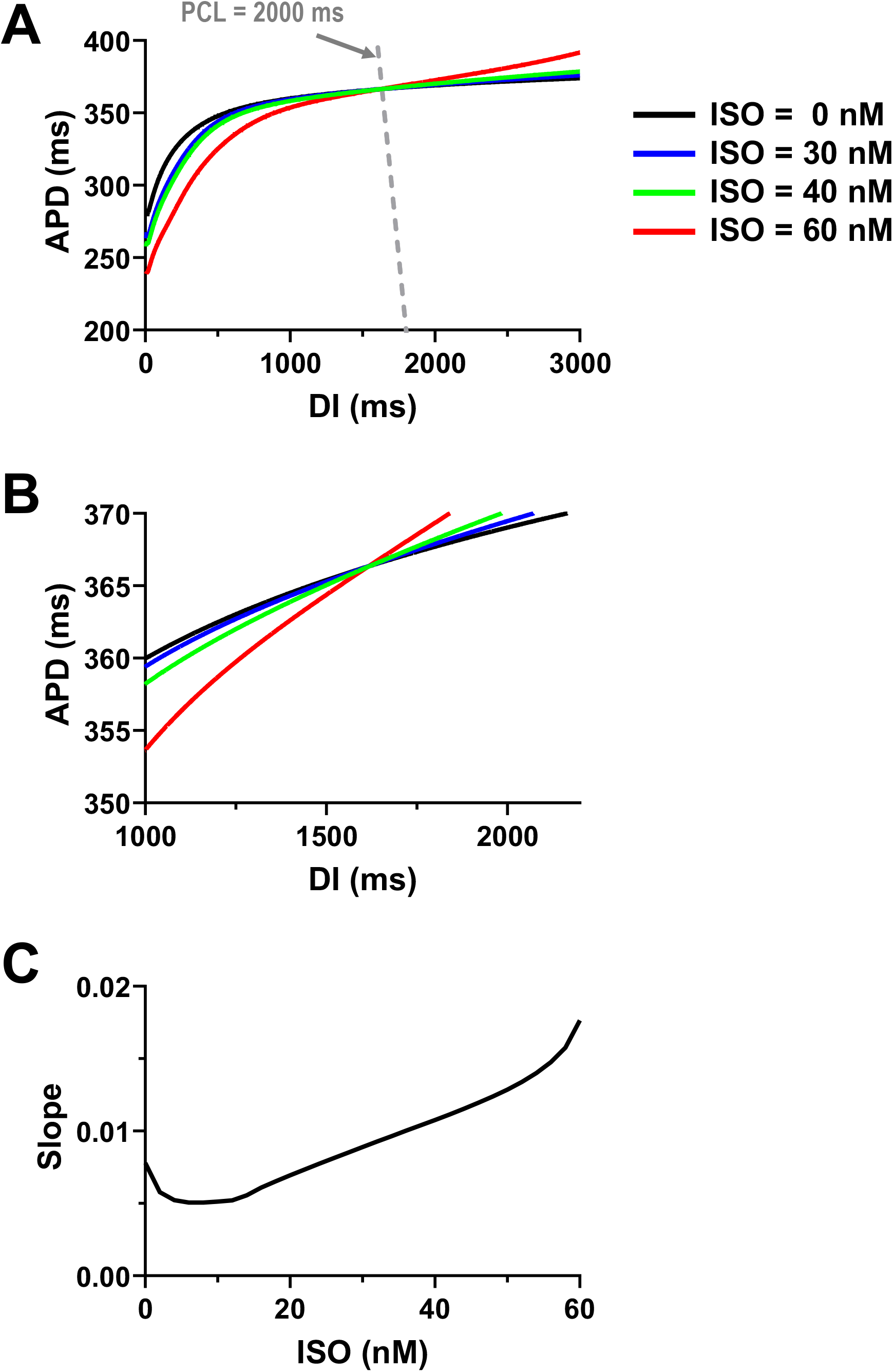
Dynamical Instability Parallels APD Variability, Not Mean APD. **(A)** S1-S2 APD restitution curves under constant mean APD conditions. The gray dashed line indicates PCL = 2000 ms (PCL = APD + DI). The restitution slope is evaluated at the intersection points of this line and the restitution curves. **(B)** Magnified view of restitution curves. The plots illustrate that while the APD at PCL = 2000 ms is identical, the restitution slopes at that point differ between ISO concentrations. **(C)** The restitution slope as a function of ISO concentration. The profile of the slope (instability) parallels APD variability (Fig. 4C), but not the mean APD (Fig. 4B).

## Discussion

### Dissociation of Repolarization Duration and Stability under β-Adrenergic Stress

The central finding of this study is the dissociation between the duration and dynamic stability of the cardiac action potential under simulated β-adrenergic stimulation. Our computational model demonstrates that while increasing concentrations of ISO progressively shorten APD, a response conventionally interpreted as benign or antiarrhythmic, it simultaneously induces a paradoxical increase in beat-to-beat APD variability. This rise in variability parallels progressive steepening of the APD restitution slope, even under conditions in which the APD is held constant. These findings indicate that APD variability captures changes in the sensitivity of repolarization dynamics that are not apparent from mean APD alone. This enhanced sensitivity to perturbation provides a mechanistic link between APD variability and reduced repolarization stability.

### Mechanistic Link Between Cellular Instability and Clinical QT Variability

This study bridges the gap between cellular instability and clinically observable QTV. While clinical studies have long associated increased QTV with arrhythmia risk and sudden cardiac death, the underlying drivers have remained incompletely understood. Our simulations suggest that increased beat-to-beat variability arises because β-adrenergic stimulation increases the sensitivity of repolarization to stochastic perturbations. This behavior is accompanied by restitution steepening and ultimately by increased susceptibility to EAD formation. Thus, increased variability in cellular repolarization is not merely stochastic noise but reflects altered underlying electrophysiological dynamics, driven by the interplay between enhanced I_CaL_ and I_Ks_. These findings provide a potential cellular mechanism contributing to the QTV observed experimentally and clinically during enhanced sympathetic stimulation. Consequently, QTV should be viewed not simply as a statistical fluctuation, but as a robust, non-invasive metric to detect the dynamical instability of the myocardium.

Across human and animal studies, β-adrenergic activation increases both beat-to-beat APD variability and spatial APD dispersion. It also steepens APD restitution and thereby creates a more arrhythmogenic substrate. Conversely, β-blockade consistently dampens these effects. This highlights the central role of sympathetic and β-adrenergic tone in governing APD variability. For instance, sympathetic activation increases beat-to-beat APD variability in humans with heart failure. Brief sympathetic surges during the Valsalva maneuver increased the short-term variability of the activation recovery interval from approximately 3.7 to 5.3 ms. In this context, β-blockade with bisoprolol blunted this rise (32). Furthermore, acute β-adrenergic blockade in patients with normal ventricles reduced both low-frequency oscillations of APD and beat-to-beat variability, producing larger reductions in individuals with higher baseline variability (33). Finally, in a mouse myocardial infarction model, denervated infarcted hearts demonstrated greater APD_90_ dispersion than controls. These hearts also exhibited supersensitivity to the β-agonist isoproterenol, resulting in greater APD shortening and persistent dispersion. In contrast, hearts with restored sympathetic innervation did not develop this increased dispersion (34). Pathologic substrates like hypertrophic cardiomyopathy can further exacerbate these sympathetic stress responses. Recent studies demonstrate that β-adrenergic stimulation in human hypertrophic cardiomyopathy cardiomyocytes delays I_CaL_ inactivation and prolongs APD (35,36). This altered gating behavior under sympathetic tone would likely amplify the dynamical instability and beat-to-beat APD variability observed in our baseline model.

### Implications for Cardiac Safety and Regulatory Frameworks

Our findings have potential implications for contemporary approaches to cardiac safety assessment. Although hERG block and QT prolongation remain important components of proarrhythmia risk evaluation (8-11), current frameworks such as the Comprehensive in vitro Proarrhythmia Assay (CiPA) increasingly integrate multiple sources of information, including ion-channel pharmacology, cellular assays, and mechanistic computational modeling (37-39). We propose a critical refinement to the CiPA workflow that incorporates beat-to-beat variability as a standard output for both in silico modeling and hiPSC-CM assays. While current CiPA metrics (such as the qNet score) improve upon simple QT measurement, they may still overlook instability that occurs without prolongation. Integrating variability metrics into these pipelines could create a more holistic risk assessment that combines measures of duration (QT), stability (QTV), and morphology to better identify unsafe compounds and reduce the unwarranted attrition of safe therapeutics. Experimental validation across diverse pharmacological perturbations, comparison with established proarrhythmia metrics, and demonstration of predictive performance against clinically characterized compounds will be required to determine whether variability provides incremental value for cardiac safety assessment.

### Limitations and Future Directions

While our computational approach provides robust mechanistic insights, these findings are derived from in silico simulations and require experimental validation. Future studies should confirm the link between APD variability and arrhythmia in both isolated myocytes and intact tissue preparations. The simulations were performed primarily at a fixed PCL of 2000 ms, which was chosen to probe ventricular susceptibility to EAD formation. Because pacing was externally imposed, these simulations isolate the direct effects of β-adrenergic signaling on ventricular electrophysiology and do not reproduce the concurrent increase in sinus rate produced by isoproterenol in vivo. Consistent with the known rate dependence of EAD formation, overt EADs were preferentially observed at slow pacing. However, the increase in APD variability persisted at faster pacing rates (**Fig. S3A and Fig. S3B**), indicating that the central dissociation between repolarization duration and stability is not dependent on the bradycardic condition itself. The primary analyses were performed using the female ventricular model. Importantly, repeating the key simulations using the corresponding male model produced the same qualitative dissociation between mean APD and beat-to-beat variability (**Fig. S3C and Fig. S3D**), indicating that the central conclusion is not specific to the male electrophysiological substrate. Nevertheless, systematic investigation of sex-dependent responses, including potential differences in β-adrenergic signaling, remains an important direction for future study. Furthermore, as we focused on single-cell dynamics, subsequent research must extend these findings to tissue-scale simulations to evaluate how tissue properties, such as intercellular coupling, fibrosis, and ionic heterogeneity, affect these results, and test whether variability remains a robust predictor of instability at the tissue and organ levels.

## Conclusions

This study provides mechanistically grounded evidence that challenges the long-standing paradigm that relies on APD/QT prolongation as the primary biomarker of proarrhythmic risk. Beat-to-beat APD variability more accurately reflects cellular instability than mean APD. Variability increases even before the onset of arrhythmogenic EADs and thus serves as a precursor to these events. The temporal precedence of increased variability before arrhythmic events supports its utility as an early predictor of electrical instability. These findings establish variability as a more fundamental and reliable indicator of cellular proarrhythmic risk compared to traditional measures of static prolongation. Incorporating APD/QT interval variability as a critical biomarker in cardiac safety assessment and regulatory guidelines may improve proarrhythmia risk stratification, reduce the premature attrition of safe and effective therapeutics during drug development, and ultimately enhance patient safety.

## Data Availability

The source code will be freely accessible upon publication on our websites (GitHub / Zenodo).

## ABBREVIATIONS

APD: action potential duration,
Ca^2+^: calcium,
EAD: early afterdepolarization,
QTV: QT interval variability,
ISO: Isoproterenol

## Acknowledgments

Supported by Quad Fellowship by IIE (RS), NIH grants (R01HL149349 to DS, R01HL131517, R01HL170521, R01HL176651, R01HL141214, P01HL141084, R03AG086695 to EG) and American Heart Association grants (26RIRA1614813 and 26IPA1614798 to DS). This project also received funding from the European Commission Horizon Europe research and innovation programme under grant agreement 101137115 (RC) and from the Italian Ministry of University and Research (MIUR PRIN 2002 and PRIN 2022 PNRR, to RC).

## Figure Legends

**Figure S1**

**Time Course of Action Potential**. Membrane voltage vs. time at ISO = 0 nM (black) and ISO = 64 nM (red). At ISO = 64 nM, the APD in the deterministic model is 366.4 ms. However, stochastic noise induces EADs.

**Figure S2**

**Constant Mean APD Conditions (I**_**CaL**_ **adjustment). (A)** Required adjustment factor for G_CaL_ (relative to the ISO-modified G_CaL_) to maintain a constant APD of 366.4 ms across all ISO concentrations. **(B)** APD as a function of ISO concentration under G_CaL_ adjustment. The dashed line indicates the mean APD. **(C)** APD variability as a function of ISO concentration under constant mean APD conditions. **(D)** The restitution slope as a function of ISO concentration.

**Figure S3**

**Shortening of APD accompanied by increased APD variability occurs across multiple conditions and is not sex-dependent. (A)** APD as a function of ISO concentration at PCL = 1000 ms. The red dashed line indicates the mean APD. **(B)** Beat-to-beat APD variability as a function of ISO concentration at PCL = 1000 ms. **(C)** APD as a function of ISO concentration for the male action potential model (PCL = 2000 ms). The red dashed line indicates the mean APD. **(D)** Beat-to-beat APD variability as a function of ISO concentration for the male action potential model.

## References

1. Zipes, D. P., and H. J. Wellens. 1998. Sudden cardiac death. Circulation. 98(21):2334–2351, doi: 10.1161/01.cir.98.21.2334.

2. Srinivasan, N. T., and R. J. Schilling. 2018. Sudden Cardiac Death and Arrhythmias. Arrhythm Electrophysiol Rev. 7(2):111–117, doi: 10.15420/aer.2018:15:2.

3. Roden, D. M. 2008. Cellular basis of drug-induced torsades de pointes. Br J Pharmacol. 154(7):1502–1507, doi: 10.1038/bjp.2008.238.

4. Kannankeril, P., D. M. Roden, and D. Darbar. 2010. Drug-induced long QT syndrome. Pharmacol Rev. 62(4):760–781, doi: 10.1124/pr.110.003723.

5. Glassman, A. H., and J. T. Bigger, Jr. 2001. Antipsychotic drugs: prolonged QTc interval, torsade de pointes, and sudden death. Am J Psychiatry. 158(11):1774–1782, doi: 10.1176/appi.ajp.158.11.1774.

6. Yap, Y. G., and A. J. Camm. 2003. Drug induced QT prolongation and torsades de pointes. Heart. 89(11):1363–1372, doi: 10.1136/heart.89.11.1363.

7. De Ponti, F., E. Poluzzi, A. Cavalli, M. Recanatini, and N. Montanaro. 2002. Safety of nonantiarrhythmic drugs that prolong the QT interval or induce torsade de pointes: an overview. Drug Saf. 25(4):263–286, doi: 10.2165/00002018-200225040-00004.

8. Gotta, V., and B. Donner. 2025. QT interval prolongation: clinical assessment, risk factors and quantitative pharmacological considerations. J Pharmacokinet Pharmacodyn. 52(6):61, doi: 10.1007/s10928-025-10010-x.

9. Food, and A. Drug (2005). E14 Clinical Evaluation of QT/QTc Interval Prolongation and Proarrhythmic Potential for Non-Antiarrhythmic Drugs. U.S. Department of Health and Human Services, Food and Drug Administration.

10. ICH (2005). S7B Nonclinical Evaluation of the Potential for Delayed Ventricular Repolarization (QT Interval Prolongation) by Human Pharmaceuticals. International Council for Harmonisation.

11. ICH (2022). Questions and Answers on ICH E14 and S7B Clinical and Nonclinical Evaluation of QT/QTc Interval Prolongation and Proarrhythmic Potential. International Council for Harmonisation.

12. Vicente, J., R. Zusterzeel, L. Johannesen, J. Mason, P. Sager, V. Patel, M. K. Matta, Z. Li, J. Liu, C. Garnett, N. Stockbridge, I. Zineh, and D. G. Strauss. 2018. Mechanistic Model-Informed Proarrhythmic Risk Assessment of Drugs: Review of the “CiPA” Initiative and Design of a Prospective Clinical Validation Study. Clinical Pharmacology & Therapeutics. 103(1):54–66, doi: 10.1002/cpt.896, https://ascpt.onlinelibrary.wiley.com/doi/abs/10.1002/cpt.896.

13. Li, Z., B. J. Ridder, X. Han, W. W. Wu, J. Sheng, P. N. Tran, M. Wu, A. Randolph, R. H. Johnstone, G. R. Mirams, Y. Kuryshev, J. Kramer, C. Wu, W. J. Crumb, Jr., and D. G. Strauss. 2019. Assessment of an In Silico Mechanistic Model for Proarrhythmia Risk Prediction Under the CiPA Initiative. Clin Pharmacol Ther. 105(2):466–475, doi: 10.1002/cpt.1184.

14. Li, Z., G. R. Mirams, T. Yoshinaga, B. J. Ridder, X. Han, J. E. Chen, N. L. Stockbridge, T. A. Wisialowski, B. Damiano, S. Severi, P. Morissette, P. R. Kowey, M. Holbrook, G. Smith, R. L. Rasmusson, M. Liu, Z. Song, Z. Qu, D. J. Leishman, J. Steidl-Nichols, B. Rodriguez, A. Bueno-Orovio, X. Zhou, E. Passini, A. G. Edwards, S. Morotti, H. Ni, E. Grandi, C. E. Clancy, J. Vandenberg, A. Hill, M. Nakamura, T. Singer, L. Polonchuk, A. Greiter-Wilke, K. Wang, S. Nave, A. Fullerton, E. A. Sobie, M. Paci, F. Musuamba Tshinanu, and D. G. Strauss. 2020. General Principles for the Validation of Proarrhythmia Risk Prediction Models: An Extension of the CiPA In Silico Strategy. Clin Pharmacol Ther. 107(1):102–111, doi: 10.1002/cpt.1647.

15. Hohnloser, S. H., T. Klingenheben, and B. N. Singh. 1994. Amiodarone-associated proarrhythmic effects. A review with special reference to torsade de pointes tachycardia. Ann Intern Med. 121(7):529–535, doi: 10.7326/0003-4819-121-7-199410010-00009.

16. Woosley, R. L., C. W. Heise, T. Gallo, J. Tate, D. Woosley, and K. Romero. 2025. CredibleMeds QTdrugs List. https://crediblemeds.org/.

17. Sato, D., B. Hegyi, C. M. Ripplinger, and D. M. Bers. 2025. Beat-to-beat QT interval variability as a tool to detect the underlying cellular mechanisms of arrhythmias. J Physiol. doi: 10.1113/jp289051.

18. Sato, D., B. Hegyi, C. M. Ripplinger, and D. M. Bers. 2025. Role of dynamical instability in QT interval variability and early afterdepolarization propensity. Biophys J. 124(16):2768–2777, doi: 10.1016/j.bpj.2025.07.017.

19. Sato, D., B. Hegyi, C. M. Ripplinger, and D. M. Bers. 2025. Dynamical instability is a major cause of cardiac action potential variability. Biophys J. 124(7):1042–1048, doi: 10.1016/j.bpj.2025.02.007.

20. Medei, E. H., J. H. M. Nascimento, R. C. Pedrosa, L. Barcellos, M. O. Masuda, S. Sicouri, M. V. Elizari, and A. C. Campos de Carvalho. 2008. Antibodies with beta-adrenergic activity from chronic chagasic patients modulate the QT interval and M cell action potential duration. EP Europace. 10(7):868–876, doi: 10.1093/europace/eun138, https://doi.org/10.1093/europace/eun138.

21. Szentandrássy, N., V. Farkas, L. Bárándi, B. Hegyi, F. Ruzsnavszky, B. Horváth, T. Bányász, J. Magyar, I. Márton, and P.P. Nánási. 2012. Role of action potential configuration and the contribution of C^2^[a and K[currents to isoprenaline-induced changes in canine ventricular cells. Br J Pharmacol. 167(3):599–611, doi: 10.1111/j.1476-5381.2012.02015.x.

22. Shimizu, W., T. Ohe, T. Kurita, H. Takaki, N. Aihara, S. Kamakura, M. Matsuhisa, and K. Shimomura. 1991. Early afterdepolarizations induced by isoproterenol in patients with congenital long QT syndrome. Circulation. 84(5):1915–1923, doi: 10.1161/01.cir.84.5.1915.

23. Shetty, R., S. Morotti, V. Sobota, J. D. Bayer, H. Ni, and E. Grandi. 2025. Development and Clinical Validation of a Cross-Sex Translator of ECG Drug Responses. JACC Clin Electrophysiol. 11(9):2014–2027, doi: 10.1016/j.jacep.2025.05.015.

24. O’Hara, T., L. Virág, A. Varró, and Y. Rudy. 2011. Simulation of the undiseased human cardiac ventricular action potential: model formulation and experimental validation. PLoS Comput Biol. 7(5):e1002061, doi: 10.1371/journal.pcbi.1002061.

25. Grandi, E., F. S. Pasqualini, and D. M. Bers. 2010. A novel computational model of the human ventricular action potential and Ca transient. J Mol Cell Cardiol. 48(1):112–121, doi: 10.1016/j.yjmcc.2009.09.019.

26. Dutta, S., K. C. Chang, K. A. Beattie, J. Sheng, P. N. Tran, W. W. Wu, M. Wu, D. G. Strauss, T. Colatsky, and Z. Li. 2017. Optimization of an In silico Cardiac Cell Model for Proarrhythmia Risk Assessment. Frontiers in Physiology. Volume 8 - 2017, doi: 10.3389/fphys.2017.00616, https://www.frontiersin.org/journals/physiology/articles/10.3389/fphys.2017.00616 (Original Research).

27. Heijman, J., P. G. Volders, R. L. Westra, and Y. Rudy. 2011. Local control of β-adrenergic stimulation: Effects on ventricular myocyte electrophysiology and Ca(2+)-transient. J Mol Cell Cardiol. 50(5):863–871, doi: 10.1016/j.yjmcc.2011.02.007.

28. Volders, P. G., M. Stengl, J. M. van Opstal, U. Gerlach, R. L. Spätjens, J. D. Beekman, K. R. Sipido, and M. A. Vos. 2003. Probing the contribution of IKs to canine ventricular repolarization: key role for beta-adrenergic receptor stimulation. Circulation. 107(21):2753–2760, doi: 10.1161/01.Cir.0000068344.54010.B3.

29. Yeragani, V. K., R. Pohl, V. C. Jampala, R. Balon, J. Kay, and G. Igel. 2000. Effect of posture and isoproterenol on beat-to-beat heart rate and QT variability. Neuropsychobiology. 41(3):113–123, doi: 10.1159/000026642.

30. Taggart, P., P. Sutton, Z. Chalabi, M. R. Boyett, R. Simon, D. Elliott, and J. S. Gill. 2003. Effect of Adrenergic Stimulation on Action Potential Duration Restitution in Humans. Circulation. 107(2):285–289, doi: 10.1161/01.CIR.0000044941.13346.74, https://doi.org/10.1161/01.CIR.0000044941.13346.74.

31. Qin, M., T. Liu, H. Hu, T. Wang, S. Yu, and C. Huang. 2013. Effect of isoprenaline chronic stimulation on APD restitution and ventricular arrhythmogenesis. Journal of Cardiology. 61(2):162–168, doi: 10.1016/j.jjcc.2012.08.016, https://www.sciencedirect.com/science/article/pii/S0914508712002377.

32. Porter, B., M. J. Bishop, S. Claridge, J. Behar, B. J. Sieniewicz, J. Webb, J. Gould, M. O’Neill, C. Rinaldi, R. Razavi, J. S. Gill, and P. Taggart. 2017. Autonomic Modulation in Patients with Heart Failure Increases Beat-to-Beat Variability of Ventricular Action Potential Duration. Frontiers in Physiology. Volume 8 - 2017, doi: 10.3389/fphys.2017.00328, https://www.frontiersin.org/journals/physiology/articles/10.3389/fphys.2017.00328 (Original Research).

33. Duijvenboden, S. V., B. Porter, E. Pueyo, D. A. Sampedro-Puente, J. Fernandez-Bes, B. Sidhu, J. Gould, M. Orini, M. J. Bishop, B. Hanson, P. Lambiase, R. Razavi, C. A. Rinaldi, J. S. Gill, and P. Taggart. 2020. Complex Interaction Between Low-Frequency APD Oscillations and Beat-to-Beat APD Variability in Humans Is Governed by the Sympathetic Nervous System. Frontiers in Physiology. Volume 10 - 2019, doi: 10.3389/fphys.2019.01582, https://www.frontiersin.org/journals/physiology/articles/10.3389/fphys.2019.01582 (Original Research).

34. Gardner, R. T., L. Wang, B. T. Lang, J. M. Cregg, C. L. Dunbar, W. R. Woodward, J. Silver, C.M. Ripplinger, and B. A. Habecker. 2015. Targeting protein tyrosine phosphatase σ after myocardial infarction restores cardiac sympathetic innervation and prevents arrhythmias. Nature Communications. 6(1):6235, doi: 10.1038/ncomms7235, https://doi.org/10.1038/ncomms7235.

35. Coppini, R., M. Beltrami, R. Doste, A. Bueno-Orovio, C. Ferrantini, G. Vitale, J. M. Pioner, L. Santini, A. Argirò, M. Berteotti, F. Mori, N. Marchionni, P. Stefàno, E. Cerbai, C. Poggesi, and I. Olivotto. 2022. Paradoxical prolongation of QT interval during exercise in patients with hypertrophic cardiomyopathy: cellular mechanisms and implications for diastolic function. Eur Heart J Open. 2(3):oeac034, doi: 10.1093/ehjopen/oeac034.

36. Doste, R., R. Coppini, and A. Bueno-Orovio. 2022. Remodelling of potassium currents underlies arrhythmic action potential prolongation under beta-adrenergic stimulation in hypertrophic cardiomyopathy. J Mol Cell Cardiol. 172:120–131, doi: 10.1016/j.yjmcc.2022.08.361.

37. Vicente, J., R. Zusterzeel, L. Johannesen, J. Mason, P. Sager, V. Patel, M. K. Matta, Z. Li, J. Liu, C. Garnett, N. Stockbridge, I. Zineh, and D. G. Strauss. 2018. Mechanistic Model-Informed Proarrhythmic Risk Assessment of Drugs: Review of the “CiPA” Initiative and Design of a Prospective Clinical Validation Study. Clin Pharmacol Ther. 103(1):54–66, doi: 10.1002/cpt.896.

38. Colatsky, T., B. Fermini, G. Gintant, J. B. Pierson, P. Sager, Y. Sekino, D. G. Strauss, and N. Stockbridge. 2016. The Comprehensive in Vitro Proarrhythmia Assay (CiPA) initiative - Update on progress. J Pharmacol Toxicol Methods. 81:15–20, doi: 10.1016/j.vascn.2016.06.002.

39. Sager, P. T., G. Gintant, J. R. Turner, S. Pettit, and N. Stockbridge. 2014. Rechanneling the cardiac proarrhythmia safety paradigm: a meeting report from the Cardiac Safety Research Consortium. Am Heart J. 167(3):292–300, doi: 10.1016/j.ahj.2013.11.004.

