## Supplementary figures and images for "Beat-to-Beat Action Potential Duration Variability, Not Duration, Reflects β-Adrenergic Stimulation-Induced Repolarization Instability"

### Supplementary Figures

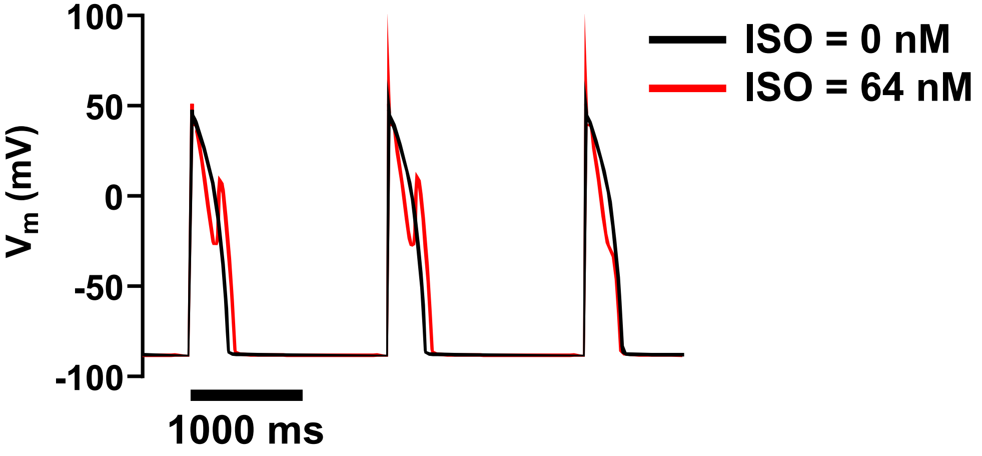


#### Figure S1

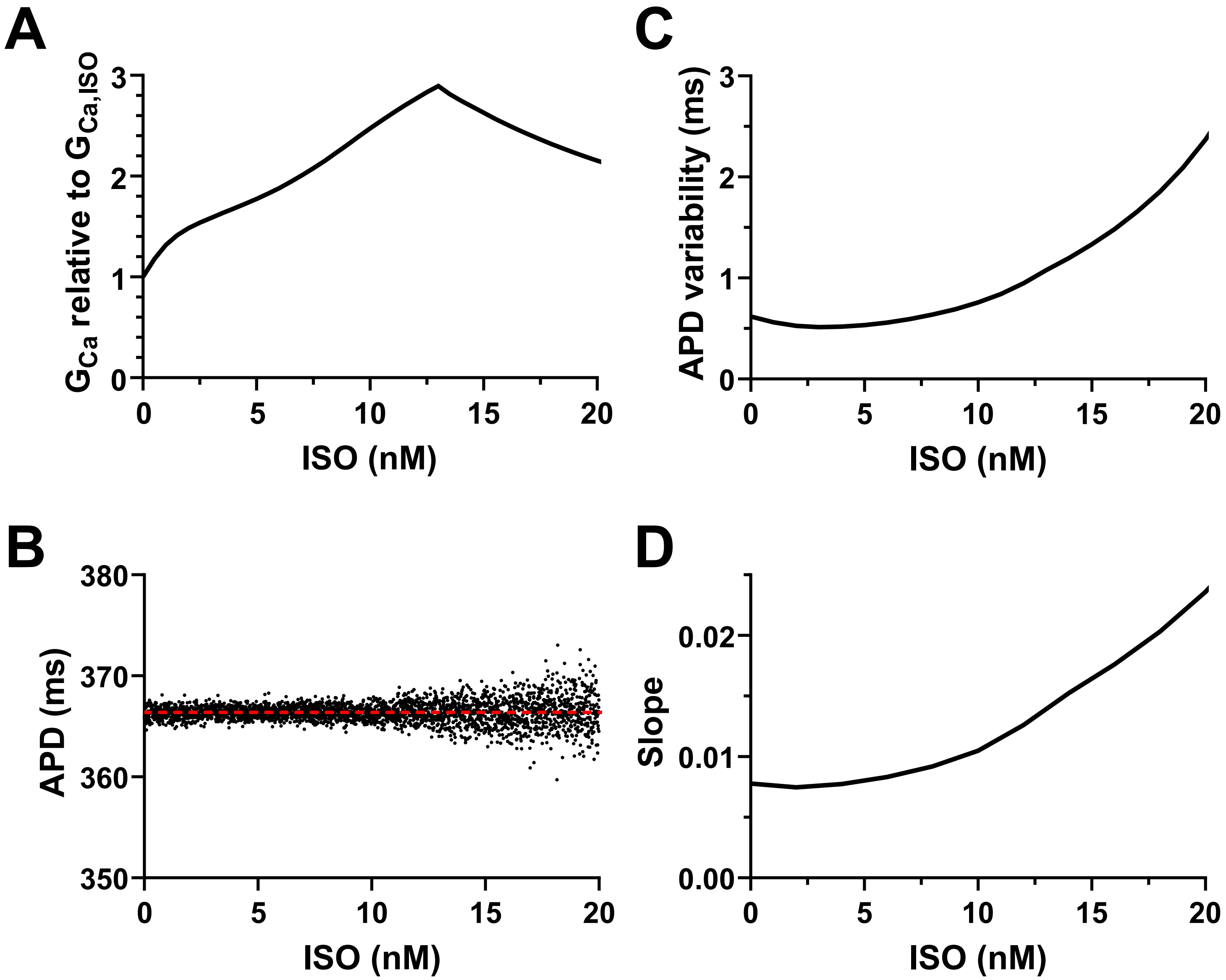

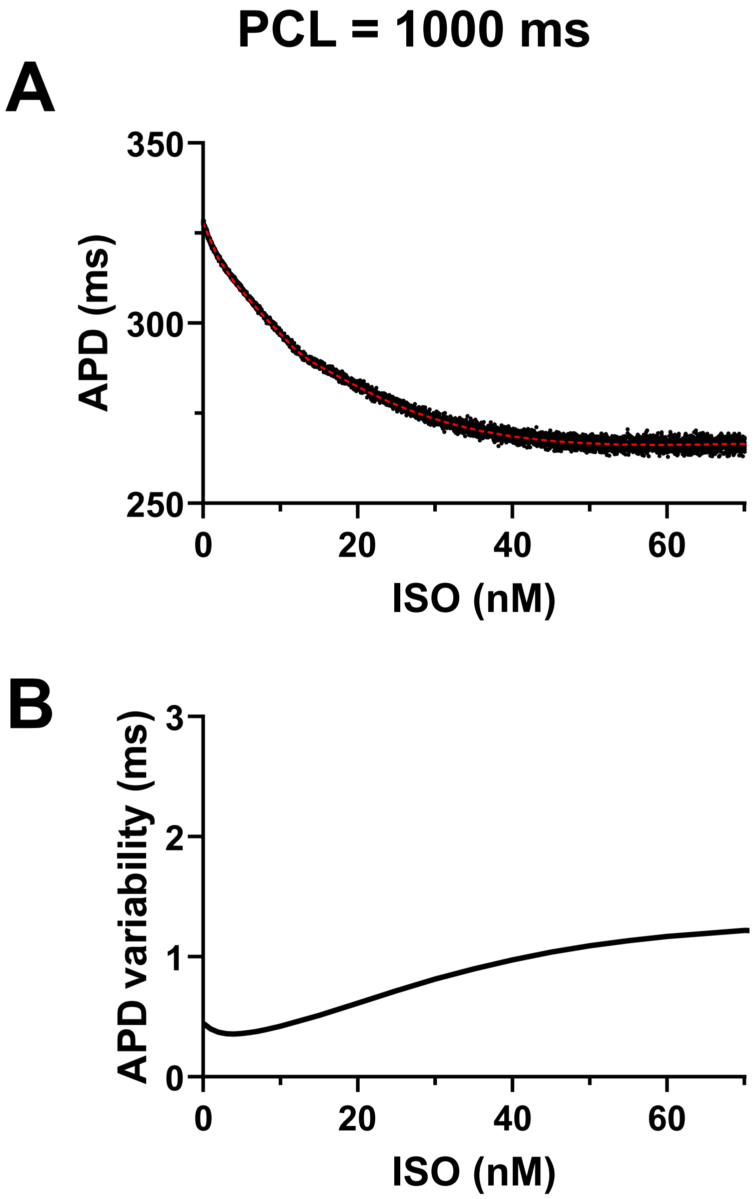

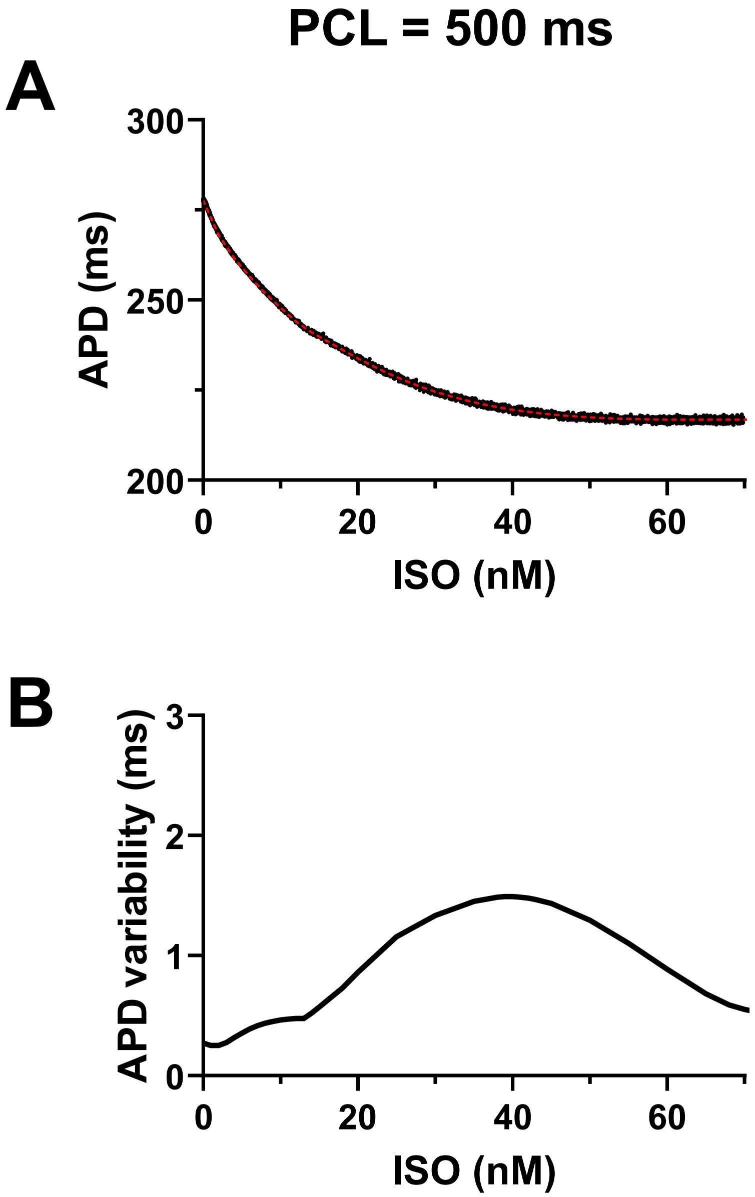
